# The lockdown of Hubei Province causing different transmission dynamics of the novel coronavirus (2019-nCoV) in Wuhan and Beijing

**DOI:** 10.1101/2020.02.09.20021477

**Authors:** Xinhai Li, Xumao Zhao, Yuehua Sun

## Abstract

**Background:** After the outbreak of novel coronavirus (2019-nCoV) starting in late 2019, a number of researchers have reported the predicted the virus transmission dynamics. However, under the strict control policy the novel coronavirus does not spread naturally outside Hubei Province, and none of the prediction closes to the real situation.

**Methods and findings:** We used the traditional SEIR model, fully estimated the effect of control measures, to predict the virus transmission in Wuhan, the capital city of Hubei Province, and Beijing. We forecast that the outbreak of 2019-nCoV would reach its peak around March 6±10 in Wuhan and March 20±16 in Beijing, respectively. The infectious population in Beijing would be much less (only 0.3%) than those in Wuhan at the peak of this transmission wave. The number of confirmed cases in cities inside Hubei Province grow exponentially, whereas those in cities outside the province increase linearly.

**Conclusions:** The unprecedented province lockdown substantially suspends the national and global outbreak of 2019-nCoV.

## 1. Introduction

A novel coronavirus (2019-nCoV) appeared in December 2019 in Wuhan, Hubei Province in central China had triggered city closure on Jan. 23, 2020, and lockdown of all major cities in the province a few days later (Fig. 1). At present, over 50 million people are constrained locally. Due to the threat of 2019-nCoV to public health, World Health Organization (WHO) declared novel coronavirus (2019-nCoV) outbreak to be “public health emergency of international concern” on Jan. 30, 2020 [1]

**Fig. 1.**
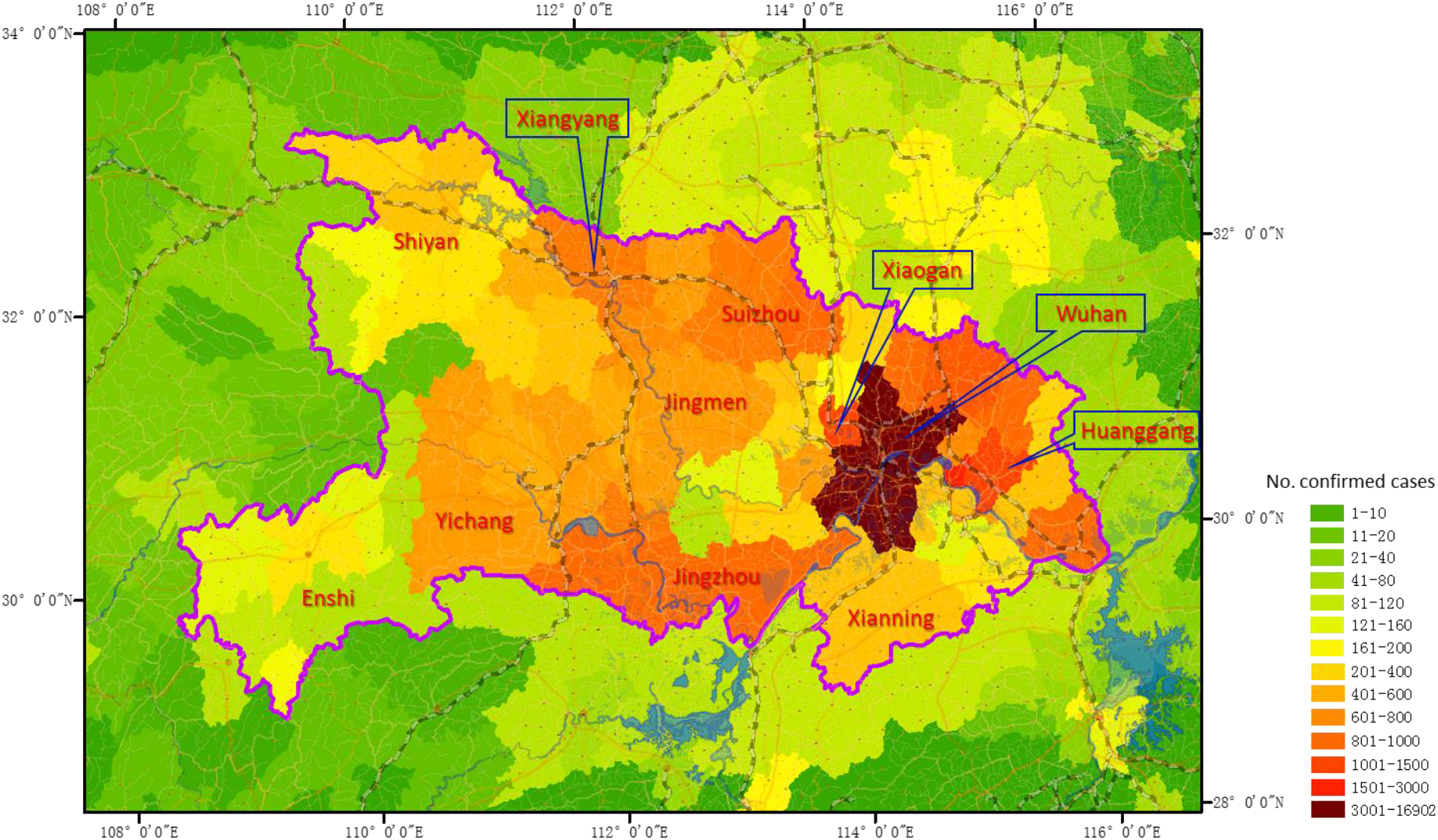
Lockdown of Hubei Province to enclose novel coronavirus (2019-nCoV). The gradient color represents the number of confirmed cases in Hubei Province and adjacent areas on Feb. 9 (24:00), 2020.

It is always a great challenge to fight effectively against a pandemic [2, 3], especially when little is known about the new virus [4]. Ideally, governments, communities and medical services take rapid, effective, rational, and proportionate responses to such health emergencies; either minimalist or maximalist responses may potentially be very harmful [5, 6].

To facilitate decision making against the 2019-nCoV, researchers had predicted the transmission dynamics in different scenarios. Read et al. [7] estimated that only 5.1% (95% CI, 4.8–5.5) of infections in Wuhan were identified; ahead of 14 days, they predicted the number of infected people in Wuhan to be greater than 250 thousand on Feb. 4, 2020. Read et al. [7] suggested, before the city closure of Wuhan, that travel restrictions from and to Wuhan city are unlikely to be effective and 2019-nCoV would outbreak in Beijing, Shanghai, etc. with much larger sizes. Leung et al. [8] estimated the transmission dynamics of 2019-nCoV in six major cities in China under six scenarios: 0%, 25%, 50% transmission reduction with and without 50% mobility reduction. However, the assumption of 50% mobility reduction is much lower than the real situation. China government enforced tourism ban on Jan. 24, and carried out other control measures such as extending holidays, closing schools, cancelling meetings, suggesting a 14-day quarantine after travel. In particular, highway traffic control is strictly implemented in many cities, towns, and villages. The majority of communities in large cities such as Beijing and Shanghai have closed to visitors.

Under the circumstance of strict control measures, we forecast the transmission dynamics of Wuhan and Beijing. Wuhan is the source of 2019-nCoV, suffered a long history (two months) of virus transmission, with lack of medical resources to quarantine exposed and suspect people. Beijing is a larger city with 22 million residents, including 10 million inbound passengers after the holidays of Chinese New Year. We believe our estimation is close to the real situation and is helpful for 2019-nCoV control.

## 2. Methods

We used the Susceptible-Exposed-Infectious-Recovered (SEIR) model to estimate the dynamics of the novel coronavirus (2019-nCoV).

### 2.1. The SEIR Model

The SEIR model has the form [9]:

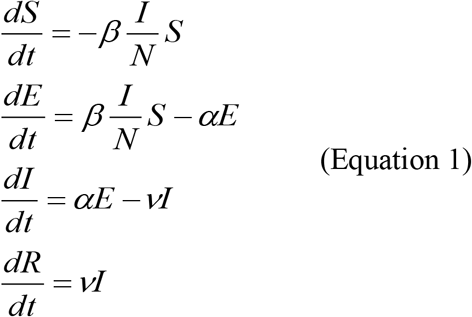

where *S* is the susceptible population, *E* is the exposed population, *I* is the infectious population, *R* is the recovered population, *t* is time (the number of days after the emergence of the first case), *N* is the total population, *β* is the average number of infected individuals per infectious subject per unit time, *α* is the reciprocal of average latent period, *v* is the rate of recovery (reciprocal of duration of the infection).

We assumed the infection rate of the novel coronavirus decreased when temperature goes up starting from March 1, 2020, since other viruses have a seasonal pattern (wave), such as the 2009 H1N1 pandemic [10]. As such, we defined the decreasing infection rate exponentially as below:

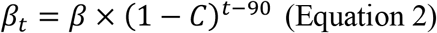

where *β*_*t*_ is the infect rate at the time *t* (the first day, *t* = 1, is Dec. 1, 2019, based on the first confirmation of 2019-nCoV on Dec. 8); *C* is a constant (simulated at 0.01-0.1) defining the decreasing rate of transmission per day.

For the cities with continuous imported infected people, we modify the growth rate of infectious population as:

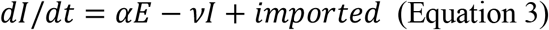

where *imported* is the daily number of imported infections.

### 2.2. Determination of Model Parameters

The basic reproductive number *R*_*0*_ for 2019-nCoV had been estimated in several independent studies (Table 1). It was noticed that changes in reporting rate of confirmed cases substantially affected *R*_*0*_ estimation [11]. In fact, *R*_*0*_ is highly associated with the intensity of control measures. We used a series values of *R*_*0*_ to fit the number of cases in Wuhan and Beijing.

**Table 1.** The estimated basic reproductive number *R*_*0*_ for 2019-nCoV

| $R_0$ | Lower CI | Upper CI | CV | Citations |
| --- | --- | --- | --- | --- |
| 3.8 | 3.6 | 4.0 | 0.05 | [7] |
| 2.68 | 2.47 | 2.86 | 0.07 | [8] |
| 2.24 | 1.96 | 2.55 | 0.13 | [11] |
| 3.58 | 2.89 | 4.39 | 0.20 | [11] |
| 2.90 | 2.32 | 3.63 | 0.22 | [12] |
| 2.92 | 2.28 | 3.67 | 0.23 | [12] |
| 6.47 | 5.71 | 7.23 | 0.12 | [13] |

The mean value of *R*_*0*_ in Table 1 is 3.51, and the mean CV (coefficient of variation) is 0.15.

The incubation period (1/*α* in SEIR model) was estimated as 6.4 (95% CI: 5.6 - 7.7) days, ranging from 2.1 to 11.1 days [14]. Liu et al. [12] provided a lower value, 4.8 days, for the period. Based on information from other coronavirus diseases, such as SARS and MERS, the incubation period of 2019-nCoV could be up to 14 days [15]. We used 6 days as the incubation period for our simulation.

Other model parameters are: population size of Beijing and Wuhan are 22 million and 10 million, respectively, based on the data (for 2018) provided by National Bureau of Statistics of China at http://data.stats.gov.cn/easyquery.htm?cn=E0105. The proportion of susceptible is 50% of the total population. The rate of recovery *ν* = 1/5 per day. The basic reproductive number *R*_*0*_ does not directly used in the SEIR model. It has the following function with the model variables and parameters: *R*_*0*_ = β/ν x S/N.

To make our analysis repeatable, we posted the data used in this study and R code for the SEIR model at https://github.com/Xinhai-Li/2019-nCoV.

## 3. Results

Until 9:00 on Feb. 9, the number of confirmed cases reaches 37251 (https://voice.baidu.com/act/newpneumonia/newpneumonia/?from=osari_pc_1). In Hubei Province, the number of confirmed cases in the capital city Wuhan and other prefecture-level cities grow exponentially; whereas in other cities in China, the number of infections increase linearly (Fig. 2).

**Fig. 2.**
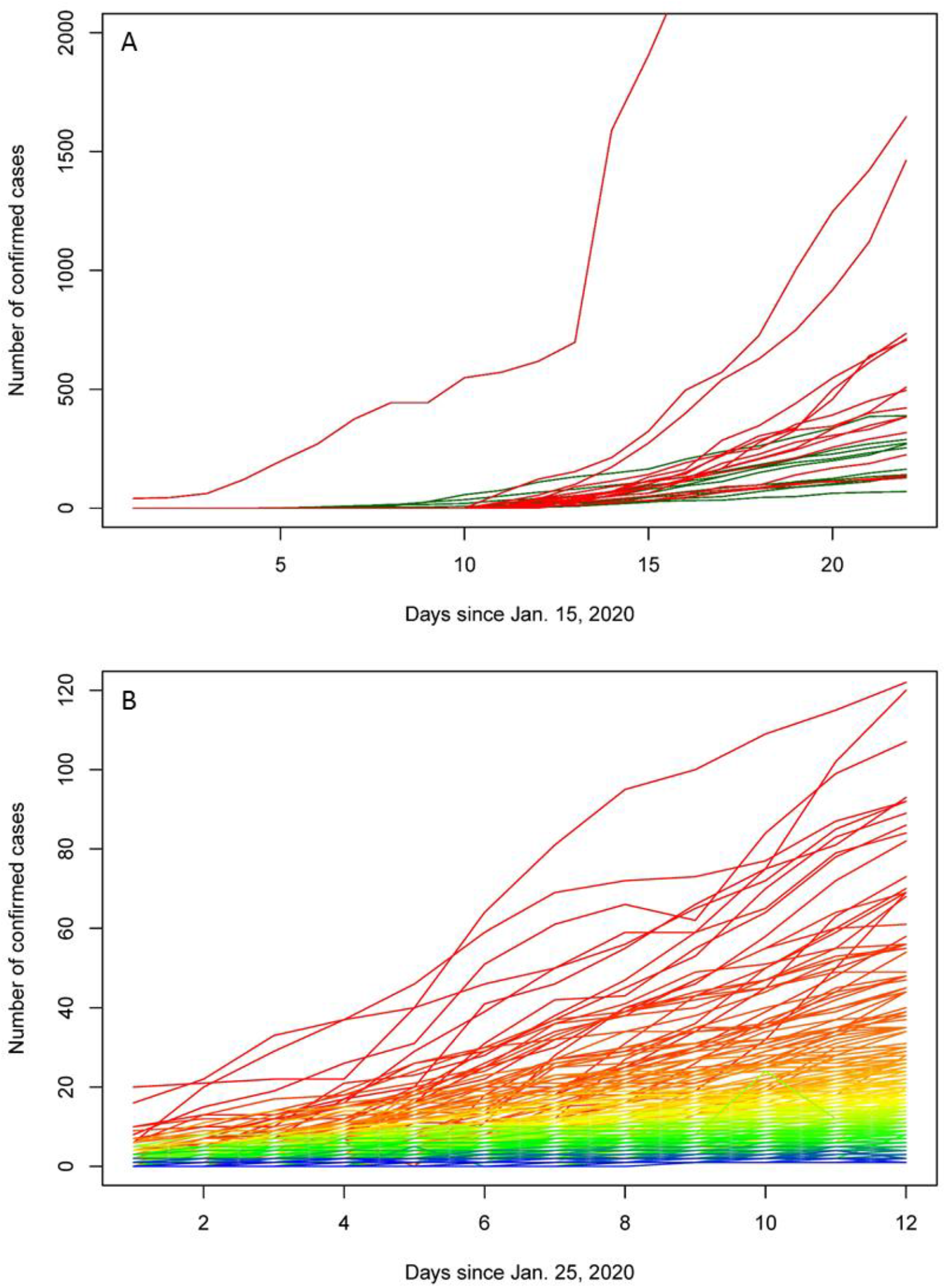
The number of confirmed cases (Jan. 25 to Feb. 5) of 2019-nCoV in 332 cities in China. A. The number of cases in top 26 cities. The red lines show the numbers of cases in cities inside Hubei Province, and the green lines demonstrate those in other cities in China. B. The number of cases in other 306 cities

We simulated the virus transmission process using the SEIR model and estimated the model parameters for Wuhan (Fig. 3) and Beijing (Fig. 4). For Wuhan, we first fit the number of confirmed cases (Fig. 3A), where the *R*_*0*_ is 5.75, much higher than that in most studies yet lower than Tang et al.’s result (Table 1). Due to the reality that a number of infected people in Hubei Province have not been checked [7, 8], we fit a more realistic transmission dynamics, under a condition of a fixed starting day on Dec. 1, 2019 (Fig. 3B). The best fit model having the *R*_*0*_ 5.0. After the city closure on Jan. 23, the residents of Wuhan performed strong self-protection by isolating themselves at home, so that we halved basic reproductive number to 2.5, reflecting the pattern of family cluster infection of the virus. Since Feb. 8, a door-to-door check was started in Wuhan in order to take all confirmed and suspected people into medical care, and the transmission between family members dropped greatly. As such, we further decrease the *R*_*0*_ to 1.5. With these transmission rates, the infectious population could reach the peak of 1.75±2.12 × 10^5^ on March 6±10, 2020. The warming climate, based on Equation 2, may shorten the transmission duration by 1-2 months (Fig. 3D).

**Fig. 3.**
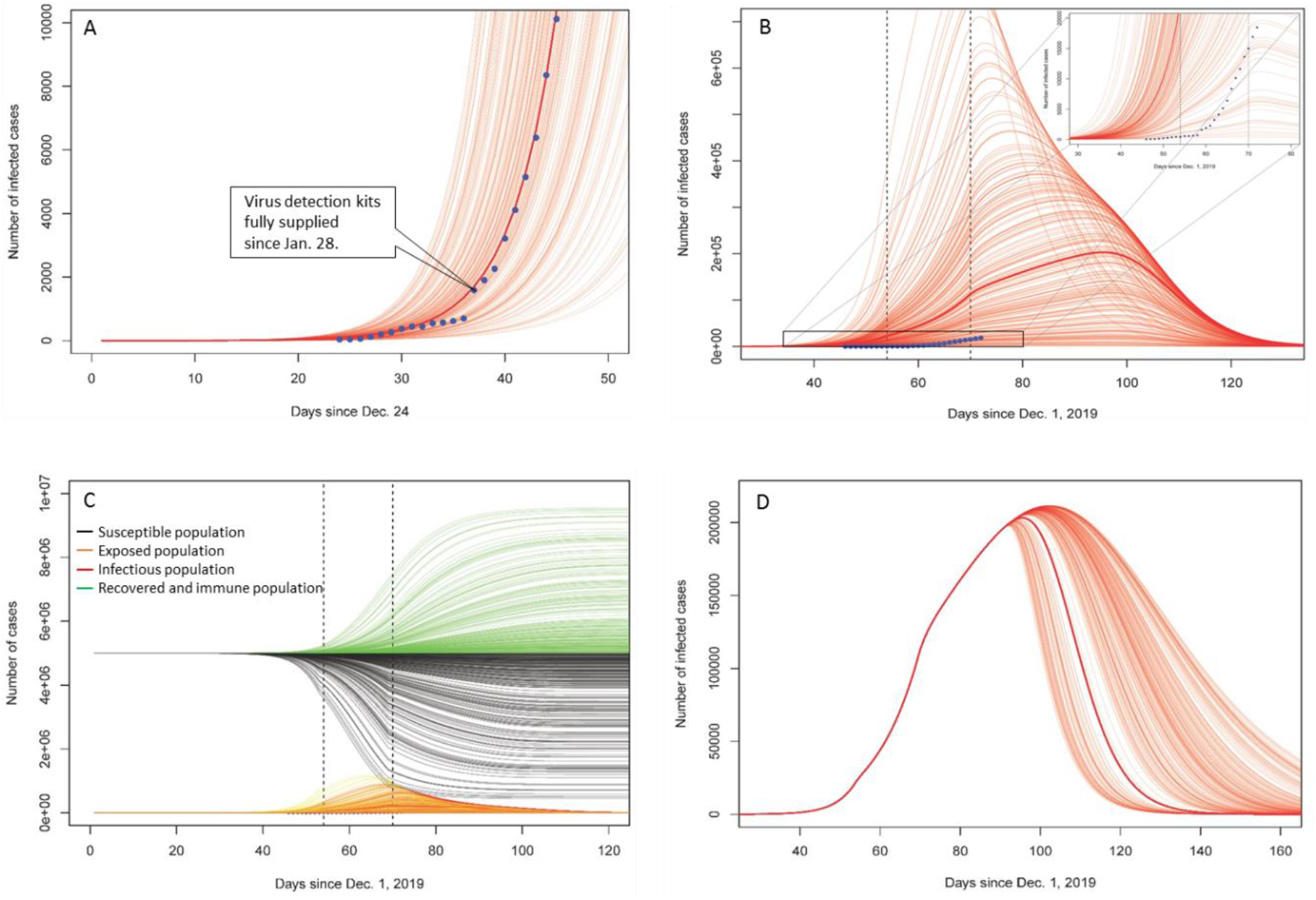
Simulated (n = 200) 2019-nCoV transmission dynamics in Wuhan. The CV of *R*_*0*_ is 15% for 200 simulations. A. Fit SEIR model to the published confirmed number of cases in Wuhan. The blue dots show the number of confirmed cases. The *R*_*0*_ is 5.75. B. Fit SEIR model based on the “real” starting date on Dec. 1, 2019. The *R*_*0*_ is 5 before Jan. 23, 2.5 between Jan. 23 and Feb. 8, 1.5 after Feb. 8. The dotted vertical lines indicate the dates of city closure (Jan. 23) and door-to-door checking (Feb. 8). C. The dynamics of susceptible, exposed, infectious and recover/immune populations. Fig. 3B is a part of Fig. 3C. D. The dynamics of infectious population under the effect of warning climate (C = 0.1, SD = 0.03).

**Fig. 4.**
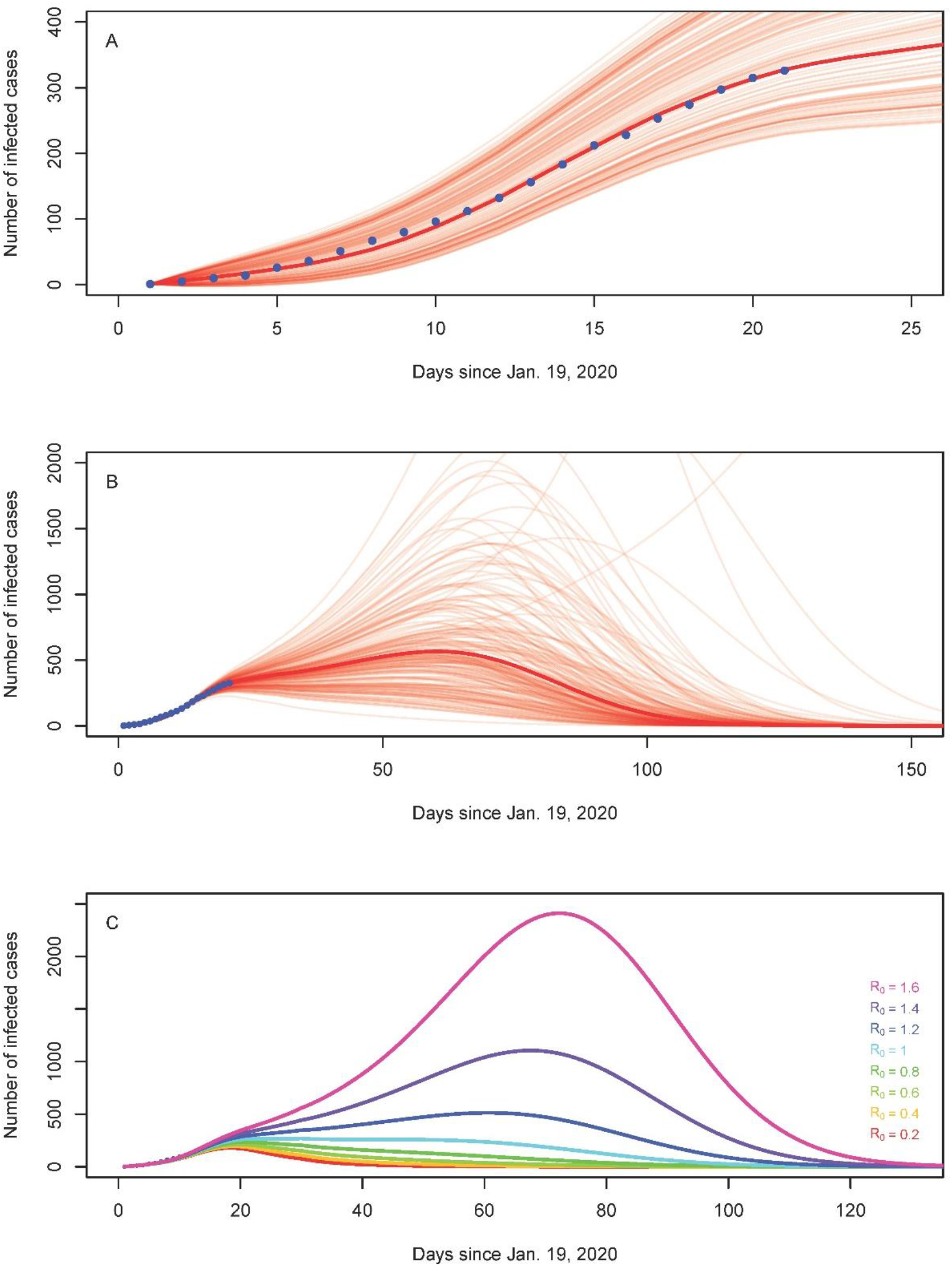
Predicted infectious population of 2019-nCoV in Beijing using SEIR. A. Predicted infectious population at the early stage of transmission. The mean imported cases of infection is 19 persons / day for 30 days, and the standard deviation of this mean value is 3.67 for 200 simulations. The *R*_*0*_ is fixed at 1.4. The blue dots show the number of confirmed cases B. Predicted infectious population by 200 simulations at the whole transmission period. The *R*_*0*_. decreases to 1.2±0.18 due to strict control. C. Predicted infectious population in Beijing with a series of *R*_*0*_ values. The parameter C (for temperature effect) in Equation 3 is 0.001 for Fig. 4B and Fig. 4C.

For Beijing, about half of the infections were directly imported from Wuhan [8]. We assume the infection flow would last for 30 days (from Jan. 19 to Feb. 18), caused by the 10 million people leaving the returning Beijing for holidays. We simulated the numbers of imported cases and found the best fit of daily imported cases for the 30 days following a normal distribution, with mean value and SD to be 19 ±12.7 individuals / day. To estimate the fluctuation of this time series of imported cases, we calculated the standard deviation of the mean value (19 individuals / day) of the daily imported patients from Jan. 19 to Feb. 8, which is 3.67. We adopted this uncertainty to run 200 simulation, and found the *R*_*0*_ 1.4 is the perfect fit (Fig. 4A). Considering the strict community locking (close to any visitors) and other extreme control measures (such as office building control, i.e. checking body temperature for every one and refusing visitors) in Beijing, we adopted a lower *R*_*0*_ 1.2 after the first 30 days of transmission. The uncertainty of *R*_*0*_ was simulated with CV = 15% (see Table 1), and the results indicate that the infectious population may reach 607±553 on March 20±16 (Fig. 4B). The uncertainty of the transmission dynamics, as simulated by the SEIR model, is very high due to large variance in imported cases and *R*_*0*_ (Fig. 4).

## 4. Discussion

We simulated the transmission dynamics of 2019-nCoV, with taking into account of the strict control measures enforced in the two cities. For Wuhan, city closure was a chock upon the local residents and they took much better protection than before. Accordingly, we halved the *R*_*0*_ after the city closure. As to Beijing, strict control measures have been implemented. As a result, the local infection happened only 41 times until Feb. 3, with 124 people infected, which equals to half of the confirmed cases [16]. The dynamics of virus transmission are dramatically different within and without Hubei Province (Fig. 2), so that we gave different model parameters for the two cities. Our results about infectious population in Wuhan is lower than Read et al.’s estimation by 60%. Our estimation for Beijing is also much lower than the prediction of other studies [e.g. 7, 8]. We believe we provide a more realistic forecast, as we are witnessing the strict prevention activities carried out by all organizations and local communities in Beijing, and the linear increase (not exponential increase) of daily confirmed cases has proved the effectiveness of control measures.

China is running an antivirus campaign against 2019-nCoV. Besides the lockdown of Hubei Province, many cities blocked the highway and stopped the traffic through their domain. The longest vacation (for Chinese New Year) had been extended twice. The majority of people quarantine themselves at home for 14 days after travel. All domestic and international tourism were cancelled and banned. Numerous meetings, games, shows have been postponed. Schools are still closed. Compared with the prevention measures against SARS 17 years ago, this campaign has much stricter control on local communities (only accessible to residents). In the meantime, every people wear a musk during outdoor activities. Mandatory quarantine for close contactors is not enforced at this time.

There are three difficulties in the antivirus campaign. 1. The incubation period of 2019-nCoV is long. 2. Some patients have no symptoms yet they are infectious. Although a case of no symptom transmission [17] had been proved flaw [18], more other cases had been reported in many sources such as newspapers, website, and Wechat, etc. The current prevention method, body temperature checking, could not detect such people. 3. The false negative rate of 2019-nCoV diagnosis is high. For example, Dr. Wenliang Li (WHO Twitter mourned his death due to 2019-nCoV on Feb. 6, 2020) had mentioned at his Wechat account that the nucleic acid detection for him was negative, when he suffered breath difficulty. We think it would have a longer fight (Fig. 3 & 4) against 2019-nCoV than SARS and MERS.

Wuhan can represent the situation of other cities in Hubei Province. After the lockdown, these cities are isolated. Even over ten thousand medical workers from other cities all over China have entered Wuhan for help, the infectious population is still too large to be taken care of. However, starting from Feb. 2 and Feb. 8, two newly established hospitals (Huoshenshan and Leishenshan Hospitals) with altogether 2500 beds for infectious patients have been already in service. With square bay hospitals having been built and coming into use, and a door to door checking of all residents in Wuhan started on Feb. 8, all infected and suspected people in Wuhan would be under medical care. The infectious population in Wuhan, under the progressive prevention activities, can be substantially lower (Fig. 3B) than the situation of natural transmission of the virus.

Beijing is a representative of other large cities outside Hubei Province in China. In Beijing, all confirmed and suspected cases have been taken into designated hospitals, with their close contactors being tracked and quarantined in hotels or at home. Similar controls are being implemented in other Chinese cities. The linear increases of total number of confirmed cases (Fig. 2) indicates the spread of the virus is under strictly control. The lockdown of Hubei Province substantially decreased the imported infections, ensure the situation being manageable outside the province.

We kept the parameters of incubation period and serial interval constant for all simulations. The uncertainty of these parameters also influences the results. The estimation of *R*_*0*_, its variance, and the rate of susceptible population, as well as the effect of control measures and warming climate, are relatively arbitrary. In spite of the weakness, we provide a more accurate estimation than previous studies. We hope our results are valuable for instructing further antivirus activities.

## Data Availability

To make our analysis repeatable, we posted all the data used in this study and R code for the SEIR model at the Github server.

https://github.com/Xinhai-Li/2019-nCoV.

